# Prenatal Exposure to the Genocide against the Tutsi in Rwanda is associated with DNA methylation at candidate genes in early adulthood: the role of trauma severity and postnatal adversity

**DOI:** 10.1101/2024.06.12.24308615

**Authors:** Luisa Maria Rivera, Glorieuse Uwizeye, Hannah Stolrow, Brock Christensen, Julienne Rutherford, Zaneta Thayer

## Abstract

We investigated associations between prenatal genocidal trauma, including maternal rape, and postnatal adverse childhood experiences (ACEs) on DNA methylation of genes associated with the stress response. In a comparative cross-sectional study of 91 Rwandan young adults, categorized by prenatal exposure to genocide and maternal rape, genocide without rape, and unexposed controls, we analyzed DNA methylation from dried blood spots and assessed ACEs and mental health at age 24. Prenatal exposure to maternal rape was associated with DNA methylation changes in *BDNF* and *SLC6A4*, with the association in *BDNF* attenuated by ACEs exposure. Genocide exposure without rape was associated with methylation changes in *PRDM8* after adjusting for early adversity. Methylation in *BDNF* and *SLC6A4* correlated with mental health scores. These findings underscore the impact of prenatal and postnatal trauma on DNA methylation and mental health, emphasizing the need for continued support for survivors in the decades after conflict.

## Introduction

The collective trauma of war can impact health and wellbeing long after the cessation of active conflict.^1^ War trauma has been associated with an increased risk of later life psychopathology, cardiovascular morbidity, and metabolic disease; these effects may be exacerbated when trauma exposure occurs during sensitive periods of development— including prenatally or during childhood. ^2–4^

The mechanisms that undergird the developmental effects of stress are complex; the shared hormonal and nutritional milieu between fetus and mother are implicated, as are the biosocial dynamics in which both mother and her child are embedded during pregnancy and childhood.^5,6^ One potential mechanism is the epigenetic regulation of neuroendocrine function. Epigenetic changes refer to chemical and/or structural changes to genetic chromatin that alter gene expression without making changes to the underlying genetic code. DNA methylation, arguably the most studied epigenetic mark, is the covalent addition of a methyl group to the 5-carbon of cytosine at cytosine-phosphate-guanine (CpG). DNA methylation can both potentiate and repress transcription, is mitotically heritable, and is largely responsible for cell differentiation, indexing early life commitments to tissue investment and neuroendocrine axis regulation.^7,8^ Previous research has shown that early life exposure to collective trauma during the Holocaust, conflicts in the Democratic Republic of Congo, Kenya, and the genocide against the Tutsi in Rwanda are associated with later life DNA methylation of genes regulating the hypothalamic pituitary adrenal axis, a core regulator of the human stress response, and multiple genes associated with metabolic function and neurodevelopment.^9–11^

While such studies examined the influence of prenatal exposure to collective trauma on DNA methylation, few account for the severity of prenatal trauma or the influence of subsequent adversity. The severity or intensity of how a trauma is experienced varies by individual, however, events that violate cultural norms (as in the case of genocidal rape) have been shown to drive subsequent distress more than “objectively” traumatizing events.^12^ Thus, assessing culturally salient forms of prenatal stress severity is critical, as more severe stress may overwhelm compensatory mechanisms that buffer the fetus from moderate stressors.^13^ Furthermore, sensitive periods of development extend beyond the prenatal period into childhood and adolescence.^14^ Epigenetic studies that examine prenatal stress should also assess postnatal experiences, as their cumulative impact may have compounding effects on epigenetic and long-term health outcomes.

The genocide against the Tutsi of 1994 elucidates the importance of these dynamics. The genocide against Tutsi resulted in the death of nearly a million Rwandans over a 100-day period of terror between April 7th and July 4th. Alongside homicidal violence, sexual assault was used as a systematic weapon of war, with an estimated 350,000 mostly Tutsi women raped during the genocide.^15^ In Rwanda, victims of genocidal rape and children who were conceived of rape have faced ongoing social stigma and ostracization in their communities, shaping access to social and material resources.^16,17^ Our previous work documents that individuals born of genocidal rape in Rwanda experience a ‘double burden’ of increased risk for childhood adversity and poorer mental and physical health relative to those conceived during the genocide without rape and controls conceived outside of Rwanda during the genocide.^18^

Literature on prenatal stress and DNA methylation has been mixed, potentially due to insufficient attention to context-specific stress severity and the mitigating or exacerbating role of postnatal experiences.^19^ To address these gaps, we conducted a study comparing DNA methylation of genes previously associated with early life adversity among individuals conceived of maternal genocidal rape (double-exposed), without maternal rape (single-exposed), and those unexposed to the 1994 genocide, while also exploring associations with subsequent adverse childhood events. Given that developmental epigenetic regulation of neuroendocrine responses is theorized to play a role in subsequent stress sensitization and mental health^20^, we also conducted an exploratory analysis of significant CpG DNA methylation and participants’ current mental health scores.

## Results

### Descriptive statistics

Participants were largely educated at the secondary level (81.3%) and categorized in either the Poor (38.5%) or Middle (52.7%) according to 2015 *Ubudehe* Rwandan government poverty categories.^36^ They reported an average of 5.6 (sd = 2.65) adverse childhood events. The mean depression T- score (48.7, sd= 10.1) and anxiety T - score (51.5, sd = 10.1) were below published screening cutoffs for depressive (≥ 53) and anxiety disorders (≥ 59)^37^, suggesting relatively good mental health in the sample (with the caveat that no clinical cutoffs have been validated in Rwandan samples to date).^22^

### Associations between genocide exposure, early life adversity, and DNA methylation in candidate genes

211 CpG sites across 17 genes were associated with genocide trauma group status (see Supplemental Table S3-S4); of these, only two survived correction for multiple testing, and only in the double-exposed vs. control contrast: cg06979684 located in *BDNF* and cg26438554 located in *SLC6A4* (see Table 2.) Following adjustment for postnatal ACEs, the site in *BDNF* no longer survived FDR, the site in *SLC6A4* remained significant, and an additional contrast between single-exposed and controls at site cg18954401 located in *PRDM8* also survived FDR. *Associations between significant CpGs and mental health* The mean Promis-29 combined depression and anxiety total raw score was 13.70 (sd =6.58) out of a possible range of 8–40, with greater scores corresponding to worse mental health. Results of the Bayesian ordinal regression indicated that methylation at the sites in *BDNF* and *SLC6A4,* but not *PRDM8,* were associated with mental health (see Table 4). A one standard deviation increase in methylation M-value at cg06979684 (*BDNF),* corresponded to a mean 2.88 increase in mental health symptoms score. At cg26438554 (*SLC6A4*), a one standard deviation increase in methylation M-value was associated with a mean score decrease of 1.14.

**Table 1.**
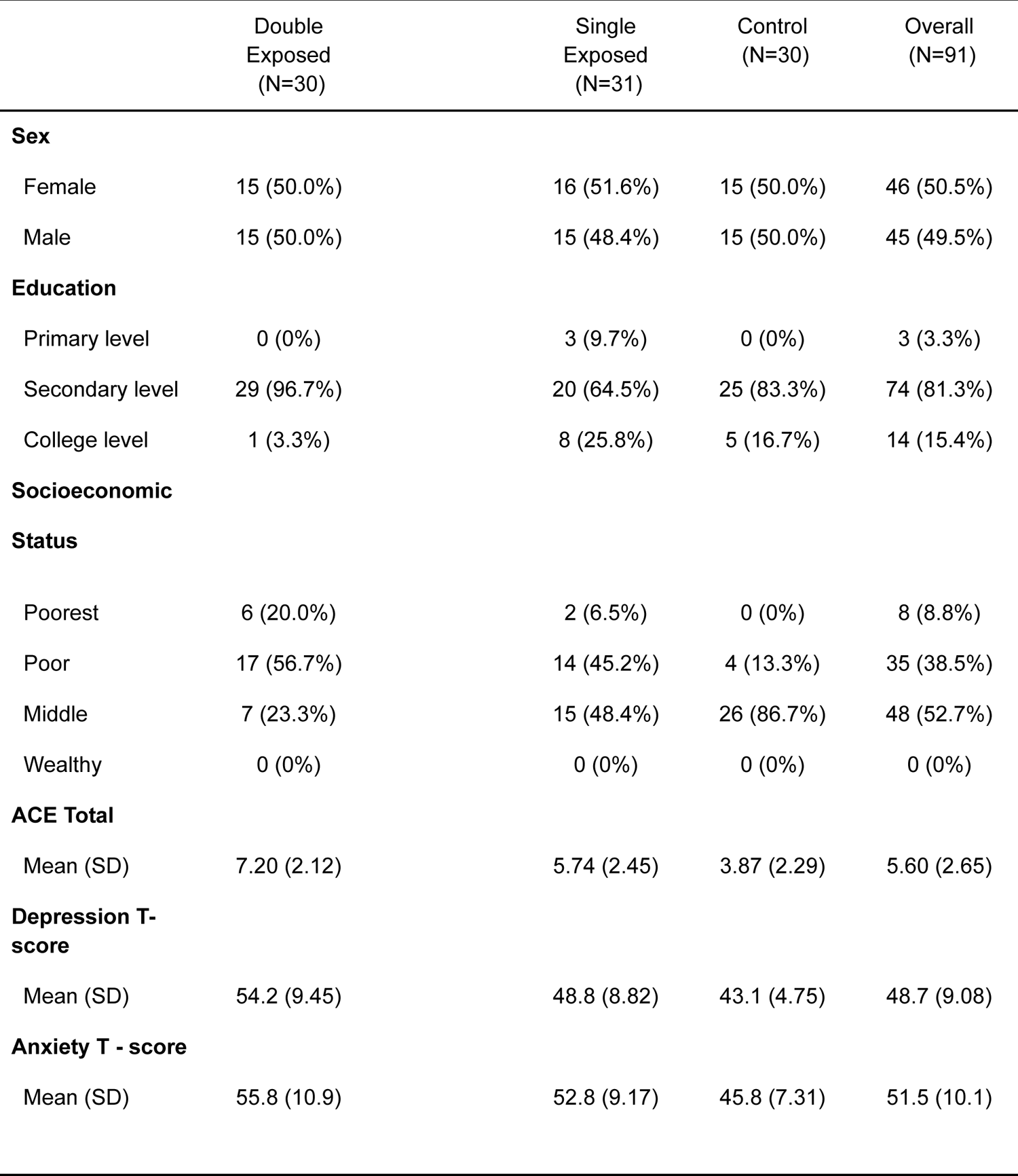
Descriptive Statistics.

|  | Double<br>Exposed<br>(N=30) | Single<br>Exposed<br>(N=31) | Control<br>(N=30) | Overall<br>(N=91) |
| --- | --- | --- | --- | --- |
| **Sex** |  |  |  |  |
| Female | 15 (50.0%) | 16 (51.6%) | 15 (50.0%) | 46 (50.5%) |
| Male | 15 (50.0%) | 15 (48.4%) | 15 (50.0%) | 45 (49.5%) |
| **Education** |  |  |  |  |
| Primary level | 0 (0%) | 3 (9.7%) | 0 (0%) | 3 (3.3%) |
| Secondary level | 29 (96.7%) | 20 (64.5%) | 25 (83.3%) | 74 (81.3%) |
| College level | 1 (3.3%) | 8 (25.8%) | 5 (16.7%) | 14 (15.4%) |
| **Socioeconomic** |  |  |  |  |
| **Status** |  |  |  |  |
| Poorest | 6 (20.0%) | 2 (6.5%) | 0 (0%) | 8 (8.8%) |
| Poor | 17 (56.7%) | 14 (45.2%) | 4 (13.3%) | 35 (38.5%) |
| Middle | 7 (23.3%) | 15 (48.4%) | 26 (86.7%) | 48 (52.7%) |
| Wealthy | 0 (0%) | 0 (0%) | 0 (0%) | 0 (0%) |
| **ACE Total** |  |  |  |  |
| Mean (SD) | 7.20 (2.12) | 5.74 (2.45) | 3.87 (2.29) | 5.60 (2.65) |
| **Depression T-**<br>**score** |  |  |  |  |
| Mean (SD) | 54.2 (9.45) | 48.8 (8.82) | 43.1 (4.75) | 48.7 (9.08) |
| **Anxiety T - score** |  |  |  |  |
| Mean (SD) | 55.8 (10.9) | 52.8 (9.17) | 45.8 (7.31) | 51.5 (10.1) |

**Table 2.** Prenatal genocide exposure ∼ DNA methylation with FDR (reference category = Control)

| Contrast | $\beta$ | SE | 95% CI | *p*-value | Probe | Gene |
| --- | --- | --- | --- | --- | --- | --- |
| Double-exposed | .22 | .055 | .12–.33 | 4.6e-05 | cg06979684 | *BDNF* |
| Double-exposed | -.33 | .075 | -.48– -.19 | 8.0e-06 | cg26438554 | *SLC6A4* |

**Table 3.**
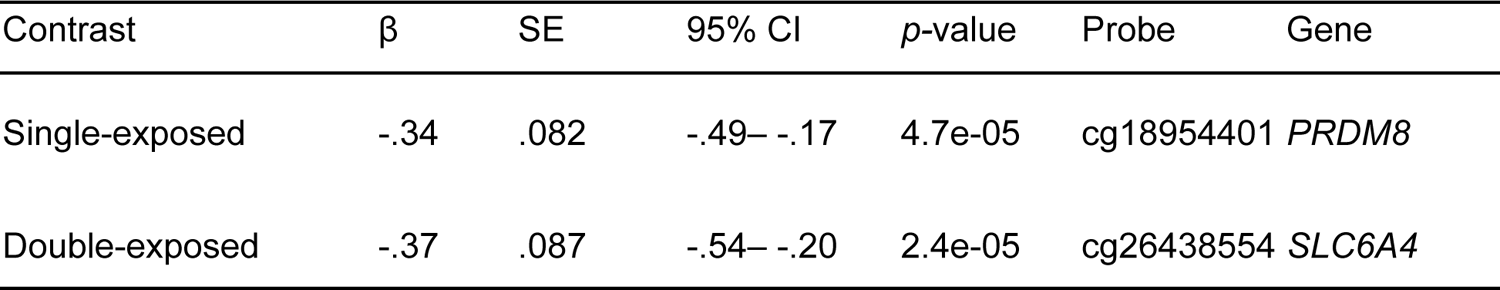
Prenatal genocide exposure + ACEs ∼ DNA methylation with FDR (reference category = Control)

**Table 4.**
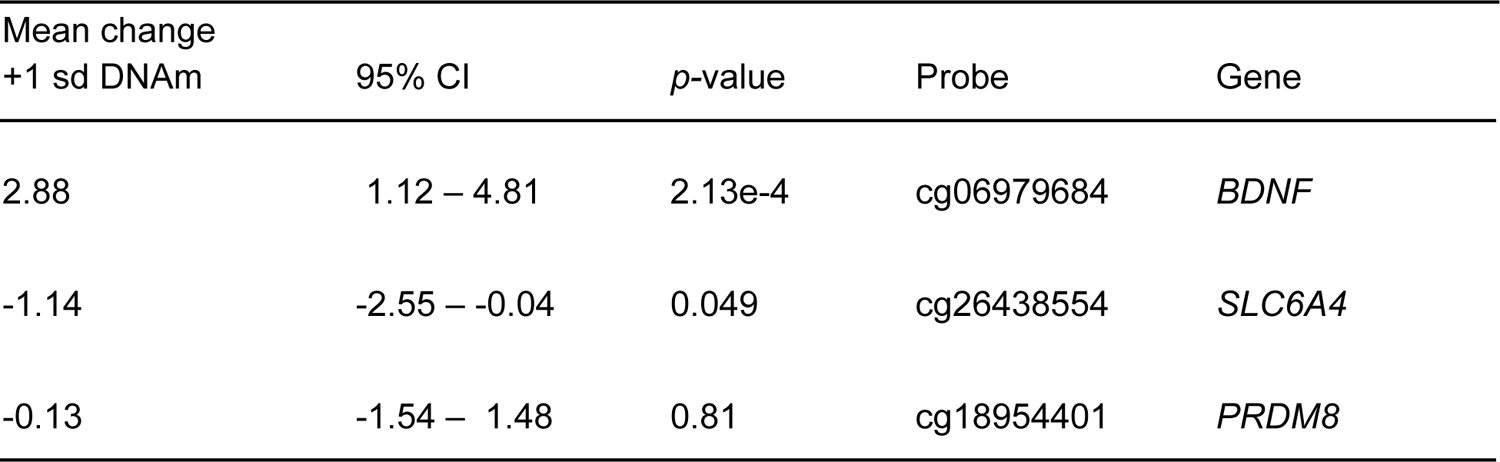
Probe specific DNA methylation and Promis-29 mental health.

| Mean change<br>+1 sd DNAm | 95% CI | *p*-value | Probe | Gene |
| --- | --- | --- | --- | --- |
| 2.88 | 1.12 – 4.81 | 2.13e-4 | cg06979684 | *BDNF* |
| -1.14 | -2.55 – -0.04 | 0.049 | cg26438554 | *SLC6A4* |
| -0.13 | -1.54 – 1.48 | 0.81 | cg18954401 | *PRDM8* |

## Discussion

We find that being conceived of genocidal rape (double-exposed) is associated with variation in DNA methylation at 2 CpG sites in *BDNF* and *SLC6A4* relative to controls after correction for multiple testing. Adjustment for postnatal ACEs attenuated the relationship at the site in *BDNF*, and revealed a new association in *PRDM8* for the single exposed. Methylation at the sites in *SLC6A4* and *BDNF*, but not *PRDM8*, were associated with mental health symptoms in the same direction as the prenatal exposure effects. In earlier work, we show that double-exposed young adults report worse physical and mental health when compared with their single exposed counterparts and controls.^18^ Our results here shed light on the physiological consequences of those original findings, providing evidence that both prenatal and postnatal stressors jointly impact adult DNA methylation, with stronger associations for those who experience more severe trauma, but which are also attributable in part to postnatal adversity.

Broadly the directionality of methylation differences, exposure status, and mental health function we found is supported by previous literature on prenatal stress and epigenetic variation. *BDNF,* which encodes brain-derived neurotrophic factor, is a well-studied neurotrophin responsible for neural differentiation, apoptosis, survival, and synaptic pruning. *BDNF* is widely expressed in the periphery and crosses the blood-brain barrier; serum levels have been explored as a transdiagnostic biomarker of mental health disorders and treatment response.^38^ Greater methylation within Exon 1 and overall lower *BDNF* transcript concentrations have been associated with major depression and anxiety disorders.^39^ Increased methylation at cg06979684, which is located in the 3’UTR in Exon1 of *BDNF*, and which may regulate transcription and splicing, has been previously found in maltreated children.^40^

Double-exposed participants had less methylation of cg26438554, located upstream of the transcription start site of *SLC6A4. SLC6A4* encodes 5 HTT, responsible for the synaptic transport of serotonin. Greater peripheral 5 HTT expression has been associated with decreased serotonin signaling and increased depressive symptoms.^41,42^ While methylation at this specific locus has not been associated with early life adversity, upstream decreased methylation of *SLC6A4* Exon 1 has previously been associated with alternative splicing of *SLC6A4*, early life adversity and major depressive disorder.^43,44^ Along with its association with more severe prenatal exposure (which did not attenuate following adjustment for ACES) we also find that decreased methylation at cg26438554 is associated with worse mental health. Further research is needed to understand whether methylation at this site and/or *SLC6A4* may be particularly sensitive to exposures during the prenatal, rather than the postnatal, period.

Finally, after adjusting for postnatal ACEs, an additional site in *PRDM8* emerges as associated with genocide exposure (without rape) contrasted with the controls. *PRDM8* encodes a histone methyltransferase responsible for modulating neuronal development and differentiation. Greater methylation of *PRDM8* has been associated with the genocide against the Tutsi in a prior epigenome-wide study that evaluated 26 individuals conceived during the genocide (16 exposed, 10 controls), but we note this study was not able to examine the severity of *in utero* maternal trauma or subsequent experiences.^28^ While no other studies specifically associate cg18954401 methylation with trauma exposure, methylation at cg18954401 is a component of the Bohlin placental clock, signalling a potential relationship to developmental conditions and less indicative of postnatal experiences.^45^

### Limitations

We note several limitations to our study. We have relatively less power to detect group differences in our small sample size and the large number of probes queried. Given our cross-sectional design with retrospective reports of early life adversity and single measure of DNA methylation, we caution against causal interpretations of our findings. Prenatal genocide trauma and maternal genocidal rape are events nested within a complex life-course socioecology, and unmeasured confounding may also explain our findings (e.g. such as SES). In addition, recruitment from community organizations may bias study participation; individuals with less access to community resources and education may have different lived and embodied experiences (although it may be argued that this may make our estimates of the impact of the genocide more conservative). Certainly, more work is needed to disentangle these factors; despite this, our replication of previous findings and use of age and sex-matched comparison groups is a study strength and supports the value of continued research.

## Conclusions

Epigenetic modifications may play a role in the increased susceptibility to mental health problems and poor health in individuals prenatally exposed to collective trauma. However, we find relationships between adult DNA methylation, prenatal trauma severity, postnatal adversity, and adult mental health are complex and should not be interpreted deterministically. In the Rwandan context, being born of genocidal rape is associated with variation in DNA methylation at two CpG sites in genes implicated in the neuroendocrine regulation of the stress response and which also showed associations with current mental health. However, adjusting for postnatal experiences attenuated the association in *BDNF* and revealed a new association in *PRDM8*, suggesting that what happens later in a child’s life has substantial impact on both their epigenome and future wellbeing, lending support to the importance of implementing systems of support in the postnatal period.

## Methods

### Participants

A sex-balanced, comparative cross-sectional sample of young adults (all 24 years of age) conceived during the genocide against the Tutsi as a result of genocidal rape (double-exposed, n= 30), conceived by genocide survivors without rape (single exposed, n=31) and conceived and gestated by Tutsi mothers outside of Rwanda during the genocide.

Institutional Review Boards of the University of Illinois at Chicago (UIC: 2018–1497), the University of Rwanda (UR No 063/CMHS IRB/2019) and Dartmouth College (STUDY0003231) approved all study procedures. Additional information on participant recruitment and enrollment in the study is available in the Supplemental Information.

### Adverse Childhood Experiences

Adverse childhood experiences were assessed by interview with a study author (G.U.), a trained mental health nurse. The ACE-IQ questionnaire assesses 13-categories of adverse childhood experiences.^21^ The ACE-IQ was translated into Kinyarwanda by a professional translator. G.U. assessed the translation and successfully piloted its use in a separate group of Rwandan young adults. Internal consistency in this sample was acceptable *(α* = 0.70). The total score was used in the analysis.

### Mental Health Promis-29

Mental health was assessed by interview with the PROMIS-29 V2.0 scale, a 29-item measure of mental and physical health concerns. Eight items assessing anxiety and depression symptoms experienced in the past seven days are rated on a 5-point Likert-like scale with higher scores indicating more severity. Widely adapted to a variety of global settings, the PROMIS-29 has strong psychometric properties.^22^ A professional Kinyarwanda translation was successfully piloted by the research team and showed good internal consistency in the present sample (*α* = 0.83). T-scores for the depression and anxiety subscales are reported in descriptive statistics to characterize the mental health functioning of the sample; the combined depression and anxiety raw subscale endorsements are used as the outcome in the ordinal regression model.

### DNA methylation

Genome-wide DNA methylation was quantified from dried capillary blood spots, which were collected from participants via sterile finger stick immediately following the interview and air-dried for 4 hours (see Supplemental Information). Two 6 mm hole punches were taken from four 125 μL spots. DNA was extracted using the QiAmp DNA Investigator Kit.^23^ DNA methylation was measured with the Illumina Infinium MethylationEPIC microarray.^24^ All samples passed a quality control pipeline using the “minfi” package in R (further described in the Supplement).

Probe-specific DNA methylation beta values were transformed to M-values to better approximate a normal distribution.^25,26^ Immune cell proportions were estimated using “FlowSorted.BloodExtended.EPIC”, which infers proportions of 12 immune cell types.^27^ We conducted a principal components analysis of immune cell types; the first component explained 68.2% of the variance and was used to adjust for cell-type heterogeneity in subsequent analyses.

### Data analysis

#### Selection of candidate genes

We focused our analysis on 19 candidate genes associated with metabolism, neurodevelopment, and the stress response that have previously been reported to be associated with early life adversity and/or war or genocide trauma exposure (see Supplemental Table S1).^11,28,29^ There were 958 total probes assessed, with 10–169 probes annotated to each gene.

#### Relationships between prenatal genocide exposure, postnatal adversity, DNA methylation and mental health

All statistical analyses were conducted in R version 4.3.3.^30^ We tested associations between prenatal genocide trauma exposure (double-exposed, single-exposed, or control), adverse childhood events, and DNA methylation in the pool of 958 probes in a three-step process using multivariable linear models and applied a Benjamini-Hochberg false discovery rate of .05 to p-values to correct for multiple testing. We first regressed probe methylation M-values onto genocide trauma group status and then repeated the analysis with the ACE total score as an additional predictor. All analyses included sex and the first cell-type PC as covariates. Sex (assigned at birth) was chosen as a covariate as it may index physiological differences relevant to DNA methylation, as well as social differences in the experience and impact of prenatal trauma exposure. Significant contrasts (with the control as the reference category) were extracted from models using the “marginaleffects” package.^31^

To investigate the potential phenotypic relevance of significant sites, we conducted an exploratory analysis of sites that survived correction for multiple testing, examining associations between methylation and the combined Promis-29 depression and anxiety endorsements.

Given the ordinal nature of the data,^32^ a Bayesian ordinal regression with a cumulative-logit link function was performed using the “brms” package in Rstan^33,34^ with random intercept for item and participant, and fixed effects for methylation M-value and sex for each of the significant CpG sites identified in the first part of the analysis. Estimates, confidence intervals, and p-value were extracted with “bayestestR”.^35^ See the Supplement for a justification of the approach and full details on the model specification.

## Data Availability

All data produced in the present study are available upon reasonable request to the authors

https://github.com/luisamariariverarivera/Rwanda_CandidateGene/blob/main/Supplemental%20Information.docx

https://github.com/luisamariariverarivera/Rwanda_CandidateGene/blob/main/Supplementary%20Tables%20S3%20%26%20S4.xlsx

## Acknowledgements

We thank the participants who shared their experiences and contributed to this study. Funding was provided by Western University Faculty of Health Sciences; Dartmouth College - Goodman Faculty Research Grant, Society of Fellows Venture Funding, Neukom Institute for Computational Science and Christensen laboratory; and University of Illinois Chicago - College of Nursing & Graduate College We thank Erik J. Ringen for his thoughtful feedback on the statistical approach.

## Author Contributions

L.M.R. conducted statistical analyses and wrote the manuscript. G.U. designed the study, collected the data, interpreted results and reviewed and edited the manuscript. Z.M.T., B.C, and J.R. contributed to study design, interpreted results, and edited the manuscript. H.S. conducted laboratory and statistical analyses and edited the manuscript. All authors read and approved the final manuscript.

## Competing Interests

The authors disclose no conflicts of interest or competing interests in this study.

## Notes

### Competing Interest Statement

The authors have declared no competing interest.

### Author Declarations

Institutional Review Boards of the University of Illinois at Chicago (UIC: 20181497), the University of Rwanda (UR No 063/CMHS IRB/2019) and Dartmouth College (STUDY0003231) approved all study procedures.

