## Supplementary Information for "Prenatal Exposure to the Genocide against the Tutsi in Rwanda is associated with DNA methylation at candidate genes in early adulthood: the role of trauma severity and postnatal adversity"

1. **Recruitment and screening of study participants**

Initial participants were selected through a convenience sampling approach from the Solidarity for the Development of Widows and Orphans to Promote Self-Sufficiency and Livelihoods (SEVOTA). SEVOTA provides aid to widows, orphans, and descendants of genocidal rape survivors. For age and gender-matched individuals born to genocide survivors who weren't subjected to rape, the initial selection was done via the Association of Genocide Widows Agahozo (AVEGA Agahozo), an organization aiding genocide survivors and their offspring. Additional participants, born to genocide survivors not affiliated with the mentioned organizations, were recruited using snowball sampling. The control group, hailing from families who had temporarily left and returned to Rwanda post-genocide, was also selected through snowball sampling in the same regions.

Many Tutsi families had left Rwanda prior to the 1994 genocide due to various factors like past political unrest in 1959, 1963, and 1973, as well as socio-economic prospects. None of the control group members originated from families who departed Rwanda due to the 1994 genocide. Participants were sourced from community groups aiding genocide survivors, and thorough screening interviews were conducted to confirm their birth connections to the genocide and rape. Efforts were made to maintain a balanced gender representation in the groups. The participants' exposure to prenatal stress related to the genocide or genocidal rape was also validated through their responses to the research questionnaires. Those conceived before or after the genocide or those with physical or cognitive impediments that could affect their participation were excluded. Data collection took place between March 7 and April 6, 2019.

2. **Rationale for selection of candidate genes**

**Table S1**

| **Gene** | **# CpG sites** | **Rationale** |  |
| --- | --- | --- | --- |
| *ADIPOQ* | 13 | Adiponectin is a cytokine secreted by adipocytes that influences insulin sensitivity and has anti-inflammatory; previously associated | Growth/metabolism |
| *BDNF* | 126 | Brain derived neurotrophic factor, regulator of neuronal development, survival and plasticity and modulator of neurotransmitter function. Previously shown to be associated with early life adversity^1^ | Neurodevelopment |
| *CRH* | 169 | Corticotropin-releasing hormone, key regulator of HPA function and the stress response. Previously shown to be associated with early life adversity.^2^ | Stress response |
| *CRHBP* | 24 | Corticotropin-releasing hormone binding protein, co-regulator of HPA function and the stress response | Stress response |
| *CYP2E1* | 26 | Cytochrome P450 2E1, membrane protein highly expressed in liver that aids in metabolic function and clearing of toxins. Previously associated with early life adversity.^3^ | Growth/metabolism |
| *FKBP5* | 48 | Glucocorticoid co-chaperone protein responsible for activation of glucocorticoid response elements. Previously associated with early life adversity and war trauma.^4^ | Stress response |
| *HOXA4* | 30 | Part of the A cluster of homeobox genes on chromosome 7 that regulate early embryonic development of the nervous system, previously associated with war trauma.^5^ | Growth/metabolism |
| *HOXA5* | 41 | Part of the A cluster of homeobox genes on chromosome 7 that regulate early embryonic development of the nervous system, previously associated with war trauma^.^^5^ | Growth/metabolism |
| *LDHC* | 10 | Lactate dehydrogenase is a key enzyme in glycolysis, previously associated with war trauma^5^. | Growth/metabolism |
| *LTA* | 61 | Lymphotoxin Alpha gene, encodes a cytokine responsible for innate immune system activation, previously associated with psychological trauma.^6^ | Immune |
| *NR3C1* | 84 | Encodes the glucocorticoid receptor, a key regulator of stress response via activation of the HPA axis and widely expressed across mammalian tissues. Multiple studies have explored prenatal exposure to stress and *NR3C1* methylation.^2,7^ | Stress response |
| *NR3C2* | 48 | Encodes the mineralcorticoid receptor, another key regulator of HPA axis function, and which mediates aldosterone impacts on salt and water balance. Implicated in prenatal stress and offspring methylation in multiple studies, including in the genocide against the Tutsi.^8,9^ | Stress response |
| *PM20D1* | 22 | Peptidase M20 Domain Containing 1 regulates hydrolase and pepsidase activity, involved in regulation of neuronal death.Previously associated with early life adversity. ^10,11^ | Neurodevelopment |
| *PRDM8* | 51 | A histone methyltransferase regulating gene implicated in epilepsy, neuronal development and differentiation. Previously associated with war trauma exposure.^5^ | Neurodevelopment |
| *SCG5* | 42 | Neuroendocrine protein 7B2 is a chaperone protein involves in neuropeptide (insulin, glucagon) maturation. | Growth/metabolism |
| *SLC43A2* | 77 | Encodes LAT4, encodes a member of the L-amino acid transporter-3 family, implicated in amino acid transport from placenta to the fetus; DNA methylation n this gene has previously been associated with early life adversity^12^. | Growth/metabolism |
| *SLC6A4* | 28 | Encodes 5HTT, the serotonin transporter, widely implicated in mental health disorders, multiple studies have found associations between methylation and depression, early life adversity, and other mental health disorder.^4,7,13,14^ | Neurodevelopment |
| *TMEM204* | 60 | Transmembrane protein 204 involved in cellular adhesion and associated with neurodevelopmental disorders. Previously associated with genocide against the Tutsi. ^5^ | Neurodevelopment |
| *VWDE* | 22 | Von Willebrand Factor D And EGF Domains, implicated in calcium ion binding and breast hypertrophy. Previously associated with genocide against the Tutsi. ^5^ | Growth/metabolism |

3. **DNA methylation extraction and QC additional methods**

*Collection of Dried Blood Spots*

Immediately following the interview, we collected dried blood spots (DBS). DBS, a minimally invasive and commonly employed technique for DNA methylation-based studies that is suitable for remote settings where it is not possible to collect and store venous blood.^15^ Whole blood drops from a sterile finger stick were collected on FTA cards (125 μL across four spots). Collection took place between March 07 and April 06, 2019. The drops were air-dried for a minimum of four hours before being enclosed in an airtight envelope containing silica-based desiccant and stored at room temperature. Subsequently, the samples were shipped from Rwanda to the University of Illinois on April 7, 2019, and later to Dartmouth College on August 26, 2021, where they were stored at -80 °C until sample processing and DNA methylation analysis in April 2022.

Quality Control Pipeline

*DNA Methylation Sample Processing and Normalization*

DNA was extracted from dried blood spots (DBS) using the QIAamp DNA Investigator Kit (Qiagen, Catalog #56504). For each of the 181 samples, two 6 mm hole punches were processed in individual 1.5 microcentrifuge tubes (Eppendorf) and QIAamp MinElute columns (Qiagen). Elution buffer ATE (Qiagen) was heated to 70°C to improve DNA release from the silica membrane. Sixty microliters of ATE were pipetted onto the MinElute column's silica membrane and incubated at room temperature (15–25°C) for 10 minutes before centrifugation. The eluate was re-eluted onto the silica membrane and incubated for an additional 3 minutes at room temperature (15–25°C). After the final centrifugation, eluates were combined into one 1.5 microcentrifuge tube, carefully mixed, and purified DNA was quantified using the Invitrogen Qubit 3.0 Fluorometer broad range assay (median = 259.6 ng of DNA). The Infinium FFPE QC and DNA Restoration kit (Illumina Inc., WG-321-1001, WG-321-1002) were utilized to assess sample quality and restore degraded DNA before bisulfite treatment (Zymo EZDNA Methylation Kit, Zymo Research, Irvine, CA, USA). DNA methylation was assessed using the MethylationEPIC v.1 beadchip, with samples randomized by the prenatal exposure group across chips.

DNA methylation microarray idat files were imported into R (version 4.2.1) and processed using the minfi package (version 1.41.0). ^16^Quality control included estimating sex and calculating mean detection p-value for CpGs across all samples to evaluate signal reliability. Beta and M-values were calculated using the normal-exponential out-of-band (Noob) method, recommended for the 12-immune-cell-type extended deconvolution, which includes normalization and background correction.^17^ Further preprocessing took place before any epigenomic analysis, including filtering CpGs with low detection p-value across samples (20,170), filtering probes on X and Y chromosomes (Y= 135, X = 18,588), and filtering SNPs/cross-hybridizing probes (77,510). DNAm Beta-values and M-values were extracted and used in subsequent analysis.

**Bayesian ordinal model of Depression and Anxiety and significant probe methylation**

Responses to the Promis29 Depression and Anxiety subscales were ordinal, rated on a Likert scale ranging from “1- Never” to “5 - Always.” Depression and anxiety subscales had 4 items each 4 items each and symptom endorsements were overall low (see STable XX) and data was heavily right-skewed and somewhat differently distributed by sex (see SFigure XX).

**Table. S2**

| **Depression** | Mean, SD |
| --- | --- |
| **Eddep04**  I felt worthless | 1.43 [0.88] |
| **Eddep06**  I felt helpless | 1.74 [1.15] |
| **Eddep29**  I felt depressed | 1.60 [0.99] |
| **Eddep41**  I felt hopeless | 1.63 [.97] |
| **Anxiety** | Mean, SD |
| **Edanx01**  I felt fearful | 1.89 [1.11] |
| **Edanx40**  I felt it was hard to concentrate on anything by my anxiety | 1.55 [0.91] |
| **Edanx41**  My worries overwhelmed me | 1.91 [1.22] |
| **Edanx53**  I felt uneasy | 1.96 [1.11] |

**Figure S1.**

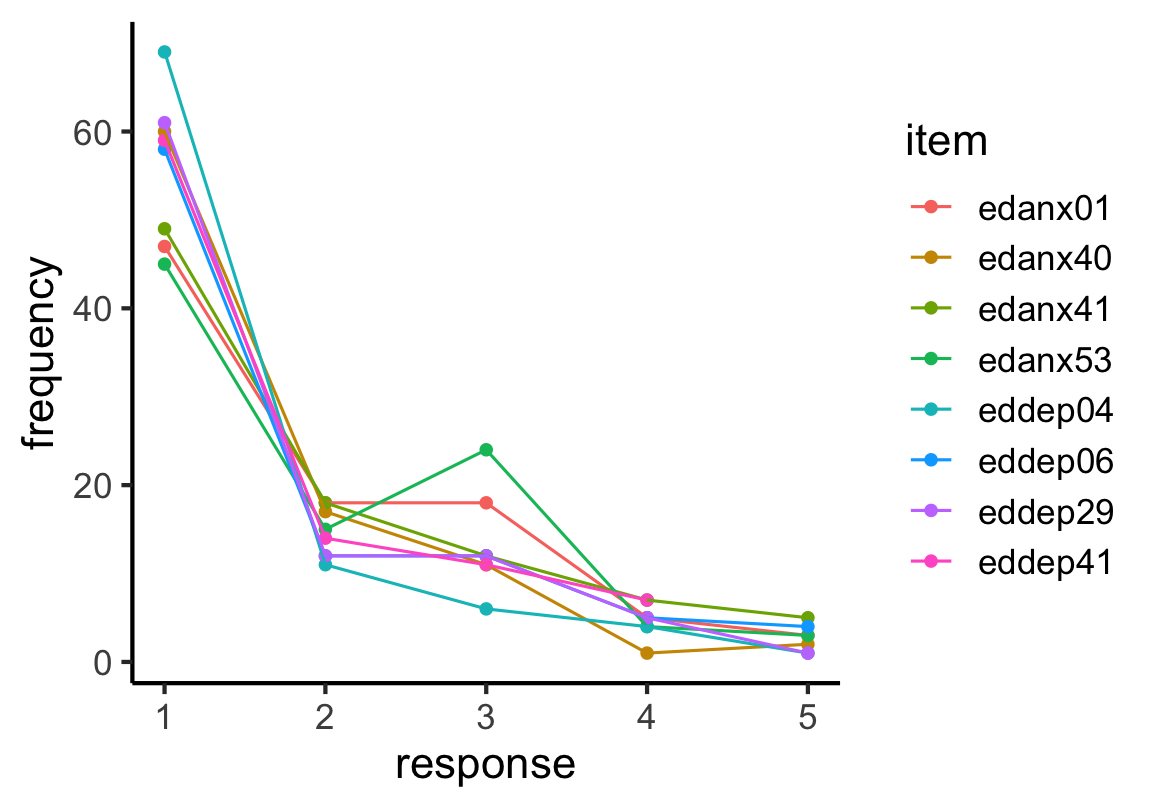

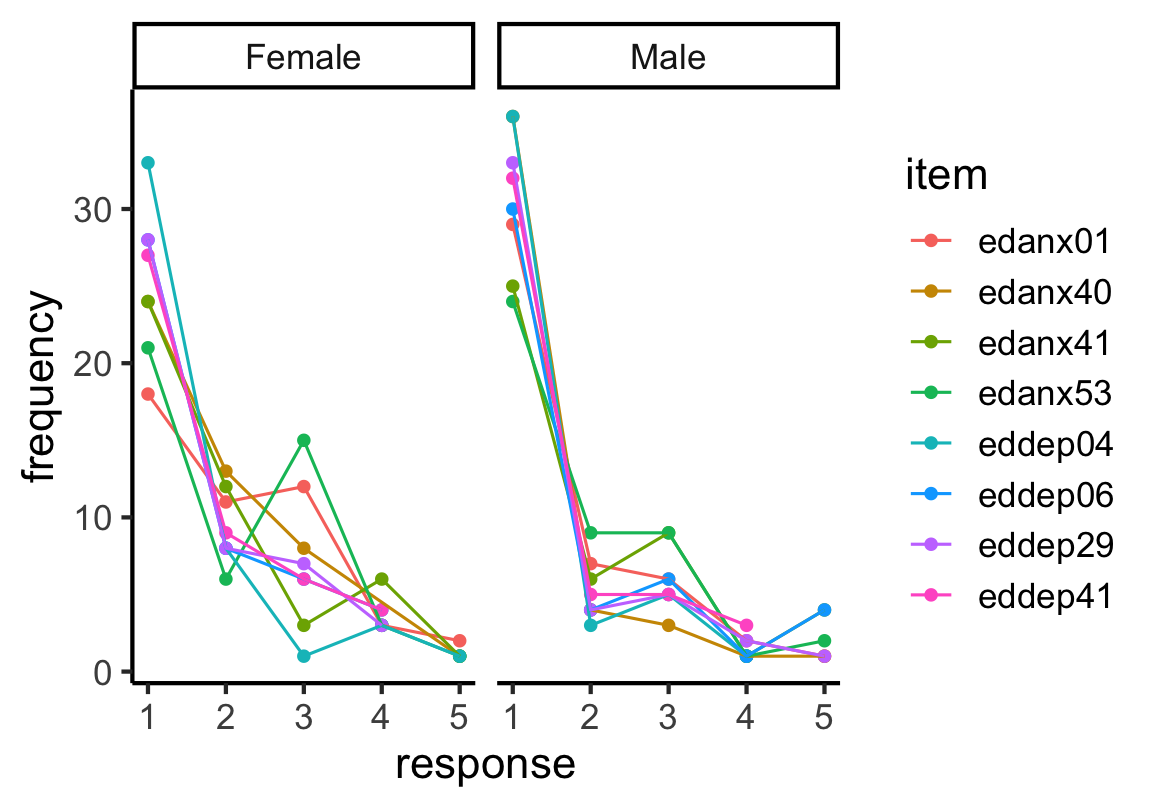

Given the low number of items per subscale, the ordinal nature of the responses, and that this was a new translation of the Promis-29 into Kinyarwanda, we believed an ordinal model would best represent our data (rather than using raw summary scores). Bayesian implementations of ordinal models are more flexible than existing frequentist implementations, and we opted to implement ours in the “brms” package in R. Our goal was to estimate how responses on the Promis-19 depression and anxiety subscales vary as a function of methylation at the significant probes identified in the first part of our analysis. Given that sex may influence both DNA methylation and responses to the Promis-29 items, we included sex as a covariate. For each significant probe, specified our model as follows:

y ~ Methylation M-value + sex + (1|item) + (1|participant)

Where M-values and sex are fixed effects, and both questionnaire item and participant id are random effects. We used generic, weakly regularizing priors for all model parameters. We monitored model converges using standard approaches (the Gelman-Rubin statistic or R-hat, effective sample size, and visual inspection of trace plots). Model performance was assessed with posterior predictive checks, contrasting model predictions against observed data. We calculated p-values and extracted model estimates and confidence intervals using the “bayestestR” package.
